# Drug Safety Warnings in English, Russian, Arabic and Hebrew Wikipedia: A Cross-Sectional Audit of 33 Drugs Across Three Risk Domains

**DOI:** 10.64898/2026.10.20.26366071

**Authors:** Maya Berlin, Matitiahu Berkovitch, Nofar Benassayag Kaduri, Einat Gorelik, Boris Gorelik

## Abstract

**Background:** Wikipedia is a leading consumer health resource, yet its medical content is audited almost only in English, and drug-safety warnings may be weaker in other editions.

**Objective:** To test whether Russian, Arabic, and Hebrew Wikipedia articles carry less actionable drug-safety warnings than English articles.

**Methods:** We audited 33 drugs in four language editions: 24 risk-positive drugs (eight each for glucose-6-phosphate dehydrogenase [G6PD] deficiency, teratogenicity, and QT-interval prolongation, benchmarked to reference standards) and nine negative controls. Three large language models, used as a measurement instrument, coded each warning 0 (not mentioned), 1 (mentioned), or 2 (explicit directive). A pharmacist resolved disagreements for G6PD and teratogenicity; the 12 QT disagreements kept the majority code, with an extreme-code sensitivity analysis. The prespecified analysis was a hierarchical Bayesian cumulative-logit model with English as reference.

**Results:** Pooled cumulative odds ratios versus English were 0.41 (Russian), 0.25 (Arabic), and 0.31 (Hebrew), with posterior probabilities of 78%, 87%, and 87% that English warns more strongly; the 95% credible intervals included 1. Eight of nine language-by-domain estimates were below 1 (exception: Russian QT). Exploratory analyses suggested that the Russian gap mainly reflects missing or short articles, whereas Arabic articles less often escalated a mentioned risk to a directive. Wikidata’s contraindication property appears in 12 statements in total, none for the audited drugs.

**Conclusions:** Warnings were weaker in Russian, Arabic, and Hebrew than in English in eight of nine comparisons, with wide uncertainty from a small purposive panel and model codes not yet validated against human coding.

**Key Points:**

- For 24 drugs with a known risk of one of three unrelated kinds of harm, Russian, Arabic and Hebrew Wikipedia articles were less likely than English articles to give readers a clear safety instruction in eight of nine comparisons, although the estimates were imprecise.
- Exploratory analyses suggest that in Russian the gap mostly reflects missing or short articles, whereas Arabic articles that mention a risk less often turn it into an instruction to avoid the drug.
- Wikidata’s dedicated contraindication field is almost unused; filling it for drugs with a known risk is a low-cost place to start closing the gap.

**Plain Language Summary:** Many people look up medicines on Wikipedia, and more and more people read its content indirectly, through AI chatbots and search summaries built on it. We checked whether Wikipedia warns readers about serious medicine risks equally well in English, Russian, Arabic, and Hebrew. We studied 33 medicines and three kinds of harm: a severe blood reaction in people with an inherited condition called G6PD deficiency, harm to an unborn baby, and a dangerous heart rhythm problem. For each article, we recorded whether it said nothing about the risk, mentioned the risk, or clearly told readers to avoid the medicine. Articles in Russian, Arabic, and Hebrew were less likely than English articles to give a clear warning, and this pattern appeared in eight of the nine combinations of language and kind of harm. Part of the gap came from articles that were missing or short. In Arabic, however, articles that mentioned a risk were still less likely to tell readers to avoid the medicine. Wikidata, the structured database that supports Wikipedia, has a dedicated field for recording the condition in which a medicine must not be used, but the field is used in only 12 places across the whole database and none of the 33 medicines has an entry, which makes it a simple place to start closing the gap.

## 1 Introduction

A contraindication protects a patient only if the person about to take the medicine can find it. Prescribers, pharmacists, and patient leaflets remain the primary sources, but many people also look a medicine up on the web, and Wikipedia is among the pages they most often reach [1,2]. Whether that page says, in the reader’s own language, that a medicine should not be used in a given condition is therefore a drug-safety question, and one that auditing English alone cannot answer.

For more than two decades Wikipedia has been among the most consulted consumer health resources: its medical articles rank at or near the top of search results and draw very large readership [1,2], and about half of physicians consult it [3]. Its direct human readership has now begun to decline as reading shifts to AI intermediaries [4,5]. The decline makes a cross-lingual audit more urgent, not less: the same articles now reach readers at one remove, as heavily up-weighted large-language-model (LLM) pretraining data [6], as the canonical grounding corpus for retrieval-augmented generation [7], and through AI search summaries that answer without sending the reader to the source [8], so what a Wikipedia edition says, and what it omits, propagates into the tools that are replacing it. This dependence is documented, not incidental: Wikipedia is among the most heavily weighted sources in LLM training corpora [9], models can reproduce its content verbatim, errors included [10], the Wikimedia Foundation itself argues that “in the AI era, Wikipedia has never been more valuable” as training data [11], and a growing list of major AI developers, including Amazon, Google, Meta, Microsoft, Mistral AI, and Perplexity, pays for structured access through Wikimedia Enterprise [12]. About one in three US adults now use an AI chatbot for health information [13,14]. These systems answer medical questions well in English but less reliably in other languages [15,16], notably Arabic [17], so an English-only view of quality is incomplete.

Fifteen years of audits find English medical content generally accurate but unevenly complete [18,19], and non-English editions are known to be shorter and less complete [20]. What has not been measured is whether the *safety-critical, patient-actionable* content (the explicit instruction not to take a drug) is equally present across languages, and whether any gap is a topic-specific accident or a systematic property of the editions.

We selected three mechanistically unrelated safety domains, each with an authoritative drug-by-drug reference standard: drug-induced acute haemolytic anaemia in glucose-6-phosphate dehydrogenase (G6PD) deficiency, one of the most prevalent enzymopathies worldwide, affecting an estimated 400–500 million people and concentrated in Mediterranean, Middle Eastern, African, and Asian populations [21,22]; drug-induced teratogenicity, whose regulatory history begins with thalidomide and more than 10,000 birth defects [23], benchmarked to current FDA pregnancy labelling [24]; and drug-induced QT-interval prolongation / Torsades de Pointes, a cause of sudden cardiac death from everyday drug classes such as antibiotics, antidepressants, and antiemetics, benchmarked to the CredibleMeds Known-Risk list [25].

Choosing domains that share no biological mechanism allowed us to examine whether cross-lingual gaps extend beyond a single safety topic.

English is the reference edition: it is the largest and most developed, and it is the language of the reference standards used here (CPIC, FDA labelling, CredibleMeds). Russian, Arabic, and Hebrew were chosen at the proposal stage of the study for their relevance to patients in Israel, where the clinical authors work, and because they serve populations in which several of the audited risks are common, including the Mediterranean basin, the Middle East, North Africa, the Caucasus, and Central Asia. They also differ in how developed the audited articles are (median length 1149 words in Russian against 490 in Arabic and 485 in Hebrew; Table 1), so the gap can be examined across editions whose audited articles differ in length. Vernacular editions matter beyond their home regions: limited proficiency in a country’s majority language is associated with lower health literacy and worse health status [26], and where subscription drug references are paywalled, Wikipedia often serves as the de facto public reference [3]. The panel is purposive and covers four of roughly 300 language editions. It tests whether a gap exists in editions that matter to these patients; it is not a sample of Wikipedia and does not support estimates for editions in general.

**Table 1.** Article coverage and structural characteristics by Wikipedia language edition (33-drug panel; medians over existing articles; the citation-density column is the median of per-article densities, not the ratio of the two preceding medians).

| Edition | Articles present | Median words | Median references | Refs / 1000 words |
| --- | --- | --- | --- | --- |
| English | 33/33 | 1989 | 105 | 51.5 |
| Russian | 26/33 | 1149 | 27 | 20.4 |
| Arabic | 32/33 | 490 | 10.5 | 20.3 |
| Hebrew | 27/33 | 485 | 4 | 10.3 |

*Refs* references

This study aimed to determine whether patient-actionable drug-safety warnings are equally present across these four Wikipedia language editions, and whether observed differences are shared across safety domains or confined to individual topics. Specifically, we sought to quantify cross-lingual differences in the presence of safety information, to distinguish absence of risk coverage from failure to turn a documented risk into an explicit patient-directed warning, and to assess the contribution of structured-data (Wikidata) gaps.

## 2 Methods

### 2.1 Design

We conducted a cross-sectional observational audit of Wikipedia article text, with no interaction with editors or readers. The unit of analysis was a single cell, defined as one drug in one language edition for one safety domain. Because the study used exclusively publicly available content and involved no human participants or identifiable personal data, research ethics approval was not required. Reporting was guided by the STROBE recommendations for cross-sectional studies [27]. The study flow is shown in Supplementary Figure S5.

### 2.2 Drug list

The panel included 33 drugs. Each domain contributed eight risk-positive drugs (24 in total), selected from the corresponding authoritative reference source. A drug is risk positive for a domain when that reference standard lists it for the domain, whatever the grade: for G6PD the CPIC guideline [22], whose grades run from “use with caution” to “contraindicated”; for teratogenicity current FDA pregnancy labelling [24]; for QT the CredibleMeds Known-Risk list [25]. Nine shared negative controls were expected to carry no warning in any of the three domains. The original 25-drug, two-domain panel was reviewed and approved by the pharmacist co-author in April 2026, and the QT domain was added in June 2026 under predefined inclusion rules. The complete drug list and corresponding reference standards are provided in the Supplementary Material.

### 2.3 Article collection

We retrieved article wikitext and metadata through the MediaWiki API, preserving section structure so that the location of each warning could be recorded. For each article we computed word count, reference count, and citation density. The English, Russian, and Arabic editions cite with `<ref>` tags, whereas the Hebrew edition predominantly uses an inline note template in their place; we counted both, since tallying `<ref>` tags alone would undercount Hebrew citations to a spurious near-zero. Articles that did not exist in a given language edition were retained in the dataset and recorded as absent rather than excluded from analysis; for analyses of warning actionability, these cells were assigned a score of 0 because no warning content was available to readers.

### 2.4 Snapshot and archived revisions

The analysed text is the article file stored in the project repository (`data/raw/articles.jsonl`), whose history shows storage on 22/3/2026, 23/3/2026, and 9/6/2026; the final snapshot, stored on 9/6/2026, is the one used for the structural metrics (its Hebrew reference counts were corrected on 24/6/2026, with the article text unchanged). For each of the 118 existing articles we queried the MediaWiki revision history for the revisions made on the stored last-edit date and compared their wikitext with the stored text; all 118 matched a revision exactly. The revision identifiers and timestamps (23/10/2022 to 9/6/2026) are listed in `data/snapshot_revisions.csv`. The logged model calls date from 23/3/2026 (G6PD and teratogenicity, first 15 drugs), 29/4/2026 (G6PD and teratogenicity, the 10 drugs added after the pharmacist review of the drug list), and 9/6/2026 (QT). Between 23/3/2026 and 9/6/2026, 25 of the 54 existing articles of the first 15 drugs were edited on Wikipedia (median change in word count 0.2%, maximum 17%; `data/snapshot_ drift.csv`); we return to this under Limitations.

### 2.5 Warning coding

Each applicable article and domain pair was coded on an ordinal actionability scale: 0 when the risk was not mentioned; 1 when the risk was mentioned without an explicit clinical recommendation; and 2 when the article carried an explicit contraindication, an avoidance instruction, or an instruction not to use the drug in the relevant clinical context.

### 2.6 Coding instrument

Final codes came from a three-model consensus used as a measurement instrument: GPT-4o (OpenAI), Gemini 2.5 Flash (Google), and Grok-3 (xAI), the last replaced by its vendor-recommended successor Grok-4.3 after a mid-study deprecation. The task requires reading full drug articles in Hebrew, Arabic, and Russian, which favours large multilingual models, and drawing the three votes from separately trained model families limits correlated errors. Using LLMs as annotators with expert escalation follows evidence that they can match or exceed human annotators on text-classification tasks [28]. Each model received the full article text through a common API at temperature 0 with a domain-specific few-shot prompt that instructed it to classify only from the article text. Unanimous codes were accepted. Otherwise, in the G6PD and teratogenicity domains a pharmacist co-author assigned the final code from the text. The 12 disagreements in the QT domain were not adjudicated: each carries the majority code of the three models (every one had a two-to-one majority), which is final for this analysis; a sensitivity analysis refits the model with all 12 set to their lowest and then to their highest model code (Results). Consistent with ICMJE guidance, the models are a method, not authors. Full prompts, model versions, and the keyword-baseline comparison are in the Supplementary Material, and inter-model agreement is reported in Table 5. The codes were not validated against human coding (Limitations).

### 2.7 Consistency checks of the instrument

Two indirect checks were computed from existing outputs; neither is a validation against human coding. First, inter-model unanimity per language over the 85 risk-positive cells with an existing article (English 24, Hebrew 21, Arabic 23, Russian 17), because lower unanimity in an edition signals a less certain instrument there. Second, the legacy keyword coder of an earlier project phase (G6PD only) was compared with the final codes on the 24 G6PD risk-positive cells with an article.

### 2.8 Statistical analysis

Because the codes are ordinal, the primary analysis was a hierarchical Bayesian cumulative-logit regression [29], `code ∼ 1 + language + (1 | drug) + (language | topic)`, with English as the reference edition, fitted with weakly informative priors in bambi 0.17 [30] on PyMC 5 [31] to the 96 risk-positive cells (24 drugs in four editions); the negative controls were analysed descriptively. Partial pooling yielded, for each non-English edition, a single pooled cumulative odds ratio versus English, which we treat as the headline estimate, together with the across-domain spread of the language effect (τ). An odds ratio below 1 means that an edition was less likely than English to carry a stronger warning. We report posterior medians with 95% credible intervals, which are robust to the heavy multiplicative tails, and the posterior probability that English warns more strongly. Robustness was checked with a frequentist cluster-robust cumulative-link model [32], a prior-sensitivity sweep, a refit without the drug random effect, a paired Wilcoxon test, and Cliff’s delta [33]; details are in the Supplementary Material.

### 2.9 Additional reporting (labelled)

There was no preregistration. The hierarchical model above is the prespecified primary analysis, and its pooled odds ratios are the confirmatory estimates. Four additions are reported with their labels: a finite-population summary (the average of the three realised domain effects, beside the population-level pooled odds ratio, which extrapolates to a further domain); a sensitivity table under HalfNormal(1) and HalfNormal(0.5) priors on the across-domain SD (τ) of the language effect, beside the default HalfNormal(5.77) of the primary fit; the joint posterior probabilities that all three pooled, and all nine language-by-domain, odds ratios are below 1; and a post hoc common-effect model with domain fixed effects, `code ∼ 1 + language + topic + (1 | drug)`, specified after the primary results were seen.

### 2.10 Mechanism analyses (exploratory)

Because non-English articles were markedly shorter, we examined whether the language effect was a length artefact with two further models. First, we refitted the hierarchical model with log(word count) as a covariate, restricted to cells whose article exists, alongside a baseline refit on the *same* rows. Second, conditional on the risk being mentioned, we modelled the probability that the article escalates to an explicit directive. Article length was considered a potential mediator of the association between language edition and warning actionability rather than a conventional confounder; consequently, the length-adjusted model was interpreted as an estimate of the association conditional on article length rather than as the primary estimate of the overall language effect. Because language edition and article length were closely associated, estimates from the length-adjusted model were interpreted cautiously. These mechanism analyses were developed after the primary results were seen and are exploratory.

### 2.11 Wikidata audit

We checked whether the structured record of each drug linked it to its domain condition through Wikidata’s dedicated contraindication property (label “contraindicated in case of”, P10946) and, for teratogenicity, the “pregnancy category” property (P3489), which records legacy US and Australian letter categories. The “side effect” (P1909) and “significant drug interaction” (P769) properties were queried as context only, because the first does not express a contraindication and the second links two drugs. Links are reported for the 33 panel drugs and for the eight risk-positive drugs of each domain, and the panel was compared with two base rates, measured on 2/10/2026: all 3,763 Wikidata items that carry an ATC code, and the number of P10946 statements on the whole knowledge base. Code and structured data are available in the project repository (see Declarations, Availability of data and material).

### 2.12 Pageview data

Monthly pageviews for each audited article were retrieved from the Wikimedia REST pageviews API for July 2025 through June 2026, using the per-article endpoint with all access methods and agent=user, which excludes traffic classified as automated. Counts were tallied per language edition and reflect only articles that exist in that edition. Seven drugs analysed solely for pharmacogenetic severe adverse reactions in an earlier phase of the project were excluded, leaving the 33 drugs of this panel.

### 2.13 Use of AI tools

Beyond the three-model classification protocol described in Section 2.6, where large language models served as a measurement instrument, the authors used AI tools as assistants in the study’s execution and writing. Claude Code (Anthropic) was used to write and review the analysis pipeline and to expand the authors’ text into readable prose under their direction. Perplexity and Claude were used for literature search. Google’s NotebookLM (Gemini) was used to help process the referenced papers. All AI-assisted output was reviewed, verified, and approved by the authors, who take full responsibility for the content of the manuscript.

## 3 Results

### 3.1 Coverage and structural quality

Article coverage is near-complete in English (33/33) and Arabic (32/33) but lower in Hebrew (27/33) and Russian (26/33), with missingness concentrated in specialised G6PD drugs. English articles are also longer (median word count 1.7 times the Russian and about 4 times the Arabic and Hebrew medians) and far more densely cited than the others: the median reference count falls from 105 in English to 4 in Hebrew, and the median citation density from 51.5 to 10.3 references per 1,000 words (Table 1).

### 3.2 Reach of the audited articles

Over the 12 months to June 2026, the 33 audited drug articles drew 10,897,920 views across the four language editions (Fig. 1). Exposure was distributed very unevenly: English accounted for 81% of the total, Russian for 16%, Arabic for 1.4%, and Hebrew for 0.8%. A drug with no article in an edition offers its readers no warning at all and contributes no views to these totals, so the figures above understate rather than overstate the cross-lingual gap.

**Fig. 1.**
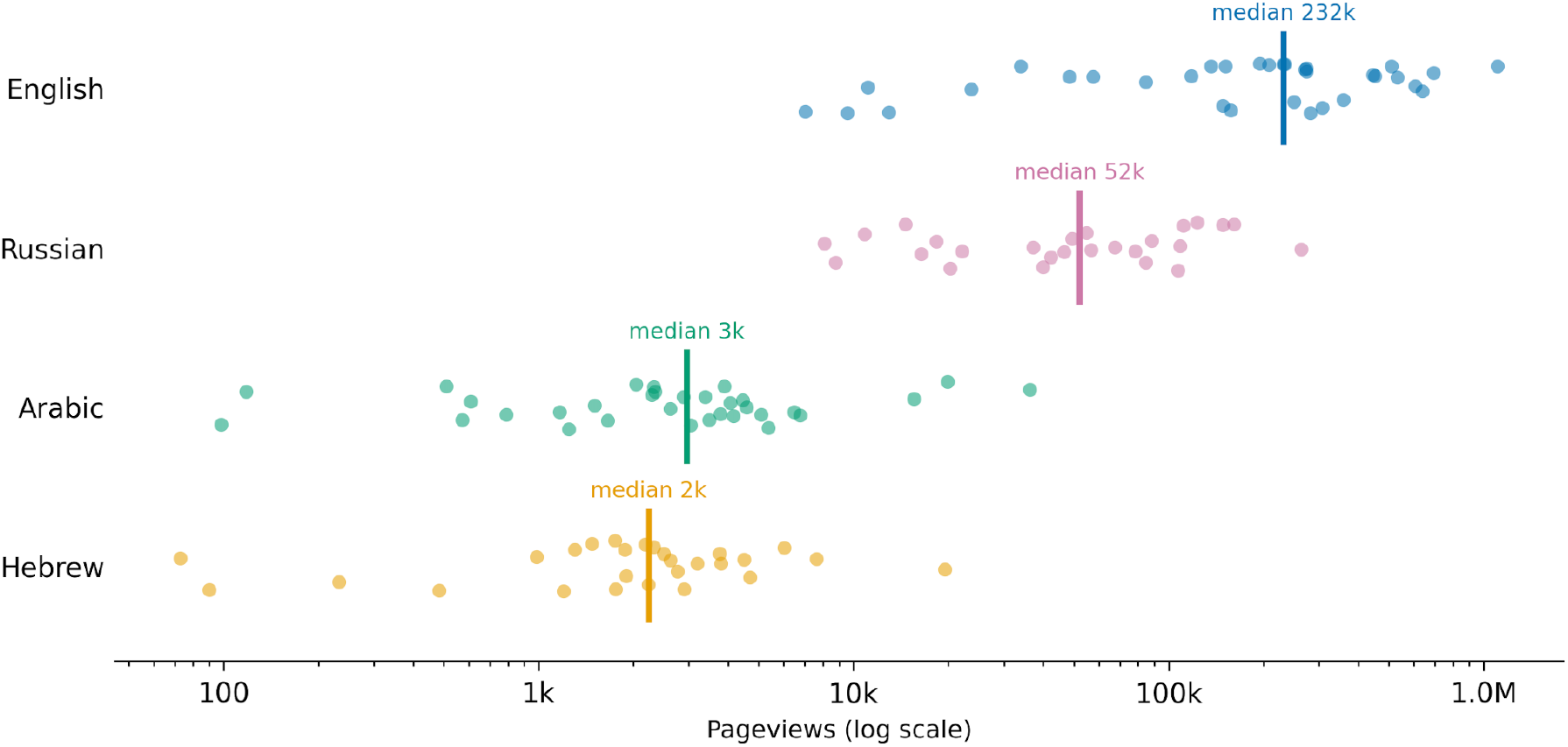
Pageviews per audited drug article by Wikipedia language edition, July 2025 to June 2026. Each dot is one drug article (English n = 33, Russian n = 26, Arabic n = 32, Hebrew n = 27); vertical rules mark the per-edition median. Editions are ordered by total views. The horizontal axis is logarithmic: the median English article drew 104 times the views of the median Hebrew article. Drugs without an article in a given edition are necessarily absent from that row. Source: Wikimedia REST pageviews API, human (non-automated) traffic

### 3.3 Warning actionability

Explicit warnings are the norm in English and the exception elsewhere, and the three domains fail differently (Fig. 2). For G6PD the non-English editions largely go silent: three-quarters of Russian and half of Arabic

**Fig. 2.**
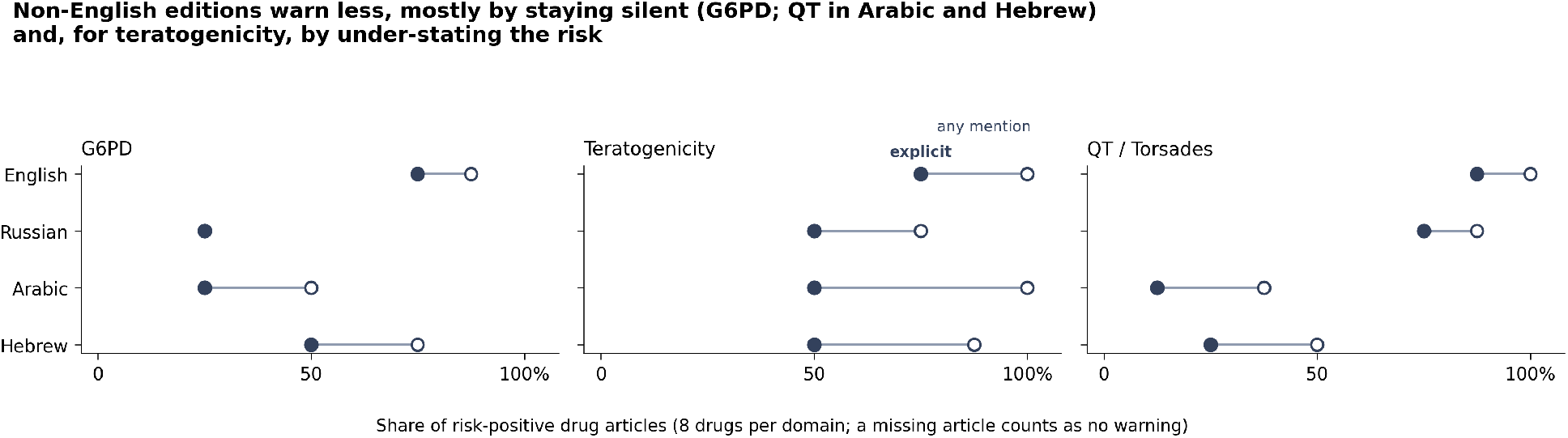
Share of risk-positive articles with any mention of the risk (open circle) versus an explicit directive (filled circle), by edition and domain (eight drugs per domain; an absent article counts as no warning). G6PD glucose-6-phosphate dehydrogenase, QT QT interval

G6PD articles do not mention the risk. For teratogenicity the risk is usually mentioned but under-stated: the share of articles with an explicit directive falls from 75% in English to 50% in each non-English edition. For QT, Arabic and Hebrew go silent (no mention in 5 of 8 and 4 of 8 articles, against none of 8 in English), whereas Russian is close to English (an explicit directive in 6 of 8 articles versus 7 of 8).

### 3.4 Confirmatory layer: direction and size of the gap

The prespecified hierarchical model quantifies the gap across all three domains (Table 2; Fig. 3). The pooled cumulative odds ratios versus English were 0.41 for Russian, 0.25 for Arabic, and 0.31 for Hebrew. All three are below 1, but the 95% credible intervals are wide (Russian 0.01 to 3.04, Arabic 0.01 to 2.10, Hebrew 0.01 to 2.15), as expected with 24 risk-positive drugs spread over nine language-by-domain cells and an across-domain SD of the language effect that the data barely constrain (τ posterior median 2.3 for Russian, 2.0 for Arabic, 1.4 for Hebrew). The posterior probabilities that English warns more strongly were 78%, 87%, and 87%. Eight of the nine language-by-domain posterior medians are below 1 (Table 2). The exception is Russian QT, at effective parity with English (OR 1.06, 95% CrI 0.01 to 7.70), and the largest and most certain gap is G6PD in Russian (OR 0.05, with a posterior probability of about 99.9% that English warns more). The joint posterior probability that all three pooled odds ratios are below 1 is 59%, and that all nine language-by-domain odds ratios are below 1 is 32%, so we describe the pattern as eight of nine estimates below 1 and not as uniform. The model converged cleanly (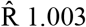, minimum ESS ≈ 1435, 0 divergences); the direction of the pooled estimates survived the robustness checks of Supplementary Appendix B, and only the magnitudes of the most extreme cells are prior-sensitive. The 12 QT inter-model disagreements carry the majority code. Refitting the model with all of them set to their lowest and then to their highest model code gave pooled odds ratios of 0.26 and 0.32 for Russian, 0.28 and 0.24 for Arabic, and 0.34 and 0.30 for Hebrew, all below 1, with posterior probabilities of 82% to 87% that English warns more (`data/coded/hierarchical_four_ topic/adjudication_sensitivity.csv`). These two extreme scenarios do not bracket the primary estimates (the Russian primary odds ratio, 0.41, lies above both), but the direction of every pooled estimate was the same under both.

**Table 2.** Language-specific differences in drug-safety warning actionability across safety domains (prespecified hierarchical model). Cumulative odds ratio versus English (posterior median; 95% credible interval for the pooled estimate). OR < 1 means less likely than English to carry a stronger warning.

| Edition | Pooled OR<br>[95% CrI] | G6PD | Teratogenicity | QT / Torsades | P(English<br>warns more) |
| --- | --- | --- | --- | --- | --- |
| Russian | 0.41 [0.01,<br>3.04] | 0.05 | 0.25 | 1.06 | 78% |
| Arabic | 0.25 [0.01,<br>2.10] | 0.11 | 0.38 | 0.04 | 87% |
| Hebrew | 0.31 [0.01,<br>2.15] | 0.33 | 0.29 | 0.09 | 87% |

*CrI* credible interval, *G6PD* glucose-6-phosphate dehydrogenase, *OR* odds ratio, *P* posterior probability, *QT / Torsades* QT-interval prolongation and torsades de pointes

**Fig. 3.**
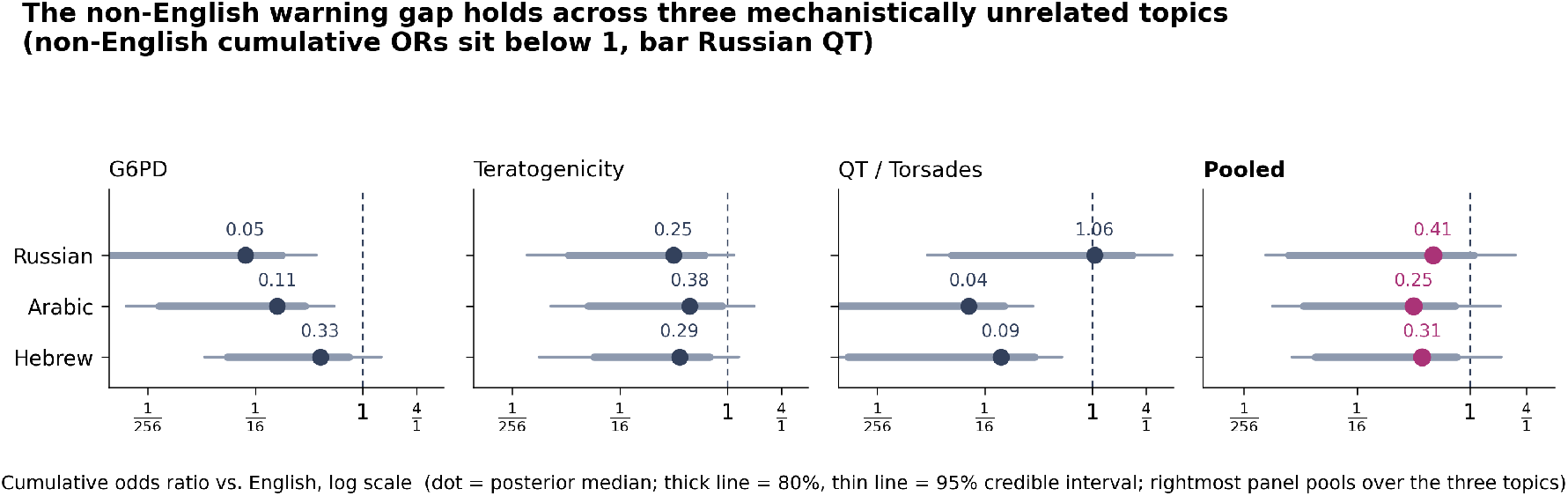
Per-domain and pooled cumulative odds ratios versus English across the three domains (posterior median, dot; 80% credible interval, thick line; 95% credible interval, thin line; dashed line = English reference). G6PD glucose-6-phosphate dehydrogenase, QT QT interval

### 3.5 Additional reporting (each labelled)

Four additions to the prespecified model (Methods) do not change the picture. (i) *Finite-population summary (descriptive complement; the prespecified estimate remains the pooled odds ratio)*. Averaging the three realised domain effects instead of extrapolating to a further domain gives odds ratios of 0.23 for Russian (95% CrI 0.02 to 0.70), 0.12 for Arabic (0.01 to 0.32), and 0.21 for Hebrew (0.03 to 0.58), each with a posterior probability of at least 98.7% of being below 1 (Table S8). (ii) *Sensitivity to the prior on τ*. Under a HalfNormal(1) prior the pooled odds ratios were 0.34 (Russian), 0.19 (Arabic), and 0.28 (Hebrew), and under HalfNormal(0.5) they were 0.33, 0.17, and 0.28, against 0.41, 0.25, and 0.31 under the default prior of the primary fit (Table S6); the intervals narrow as the prior tightens and the direction does not change. (iii) *Joint probabilities*. The joint probability that all three pooled odds ratios are below 1 is 59% under the primary prior, 84% under HalfNormal(1), and 94% under HalfNormal(0.5); for all nine cells it is 32%, 46%, and 68% (Table S7). (iv) *Post hoc common-effect model (not prespecified)*. With one language effect shared by all domains, the odds ratios were 0.30 for Russian (95% CrI 0.06 to 0.76), 0.16 for Arabic (0.04 to 0.40), and 0.25 for Hebrew (0.05 to 0.61), and the joint probability that all three are below 1 was 98% (Table S9). Its narrower intervals reflect its assumption of a shared effect as much as the data.

### 3.6 Exploratory layer: coverage gap in Russian, escalation gap in Arabic and Hebrew

These analyses were developed after the primary results (Methods). *Coverage gap*. Restricting the model to cells whose article exists, before length enters, moved the pooled Russian odds ratio from 0.41 to 0.95 (95% CrI 0.05 to 4.26), while the Arabic estimate barely moved (0.25 to 0.24) and the Hebrew estimate moved modestly (0.31 to 0.45) (Table 3). The Russian interval is wide, so the result shows that the Russian gap cannot be told apart from a coverage gap, not that no residual gap exists. Among existing articles, each e-fold of word count multiplied the odds of a stronger warning by 3.60 (95% CrI 1.35 to 7.80), and adjusting for log(word count) moved every edition toward parity, and past it for Hebrew and Russian, with wide intervals that include 1 (Table 3). Because length lies on the path from edition to warning, the adjusted estimates describe association at equal length and not the total effect. *Escalation gap*. Among cells where the risk was mentioned, Russian articles escalated to an explicit directive about as often as English ones (12 of 15 versus 19 of 23; odds ratio 1.28, 95% CrI 0.11 to 5.02), whereas Arabic articles escalated in fewer than half of such cells (7 of 15; odds ratio versus English 0.22, 95% CrI 0.02 to 0.73), and Hebrew articles showed a similar but less certain pattern (10 of 17; odds ratio 0.30, 95% CrI 0.03 to 0.99), with an interval reaching the boundary (Table 4). This comparison holds topic coverage fixed and is length-independent by construction.

**Table 3.** Exploratory. Pooled odds ratio versus English [95% CrI] restricted to existing articles, and after additionally adjusting for log(word count). Each e-fold of word count multiplies the odds of a stronger warning by 3.60 (95% CrI 1.35–7.80).

| Edition | Existing articles only | + log(word count) |
| --- | --- | --- |
| Arabic | 0.24 [0.01, 2.36] | 0.49 [0.02, 3.21] |
| Hebrew | 0.45 [0.01, 3.02] | 1.05 [0.01, 5.75] |
| Russian | 0.95 [0.05, 4.26] | 1.25 [0.01, 5.73] |

*CrI* credible interval

**Table 4.** Exploratory. Escalation from risk mention to explicit drug-safety directive across Wikipedia language editions. This comparison is length-independent by construction.

| Edition | Directive / mentioned | P(directive \| mentioned) | OR vs English [95% CrI] |
| --- | --- | --- | --- |
| English | 19/23 | 0.83 | reference |
| Russian | 12/15 | 0.80 | 1.28 [0.11, 5.02] |
| Arabic | 7/15 | 0.47 | 0.22 [0.02, 0.73] |
| Hebrew | 10/17 | 0.59 | 0.30 [0.03, 0.99] |

*CrI* credible interval, *OR* odds ratio, *P* probability

### 3.7 Warning placement

Among articles that do warn, English placed the warning in formal safety sections (contraindications or adverse effects), while the non-English editions more often carried it in running body text or the lead (Fig. 4).

**Fig. 4.**
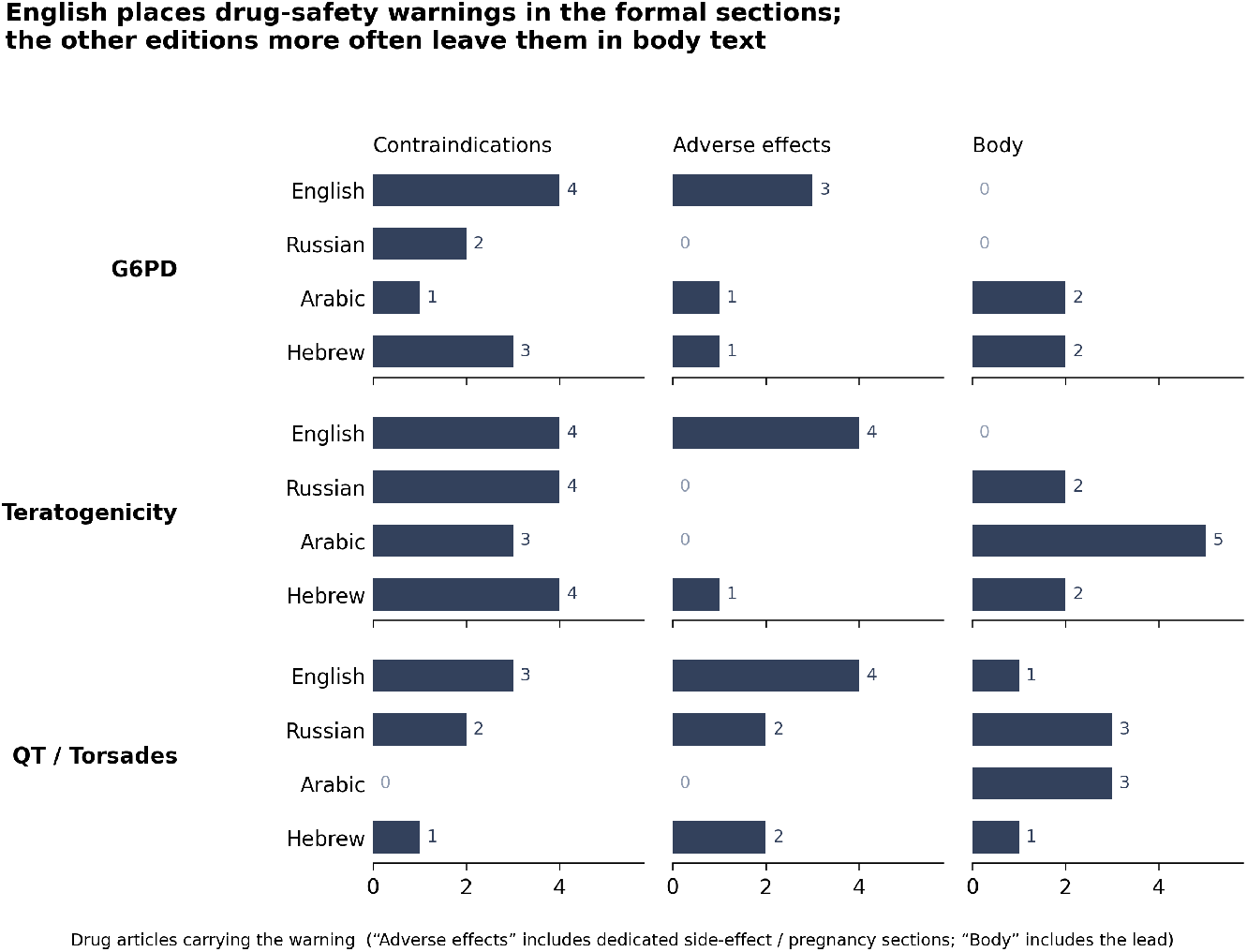
Where warnings sit, by drug group, section type, and edition. English concentrates warnings in formal safety sections (contraindications, adverse effects); the other editions more often leave them in body text. G6PD glucose-6-phosphate dehydrogenase, QT QT interval

### 3.8 Negative controls

The nine negative-control drugs were expected to score 0 in every domain and edition, giving 36 records per domain. All 36 G6PD control records were indeed 0. In the QT domain, three records were non-zero (famotidine in English; loperamide in English and Russian), each a mention without a directive; both drugs have documented QT signals at supratherapeutic doses, so these are faithful readings of the articles. In the teratogenicity domain, nine of 36 records were non-zero; on review, every one reflected pregnancy-risk language genuinely present in the article, not a coding error. Two of the nine were in English (atorvastatin and loperamide), so deviations were not confined to the non-English editions. The clearest case is atorvastatin in English, where the article still carried the explicit pregnancy contraindication that the FDA rescinded only in 2021 [34]: a regulatory lag on Wikipedia, not a language effect.

### 3.9 Structured data

Wikidata’s dedicated contraindication property (P10946) is used in only 12 statements across the whole knowledge base, and by 1 of 3,763 ATC-coded items; none of the 33 audited drugs carries it, so none of the eight risk-positive drugs in any domain (G6PD, teratogenicity, or QT) is linked to its condition through it. The zero is therefore close to the base rate, a property-adoption gap and not a drug-specific omission. Pregnancy risk is recorded for 7 of the 8 teratogens (27 of 33 panel drugs; 960 of 3,763 ATC-coded items, 25.5%), but only as legacy US and Australian pregnancy categories (P3489), not as a statement that the drug must not be used in pregnancy. The side-effect and drug-interaction properties link no panel drug to any of the three conditions, although 13 of the 33 panel drugs carry at least one drug-drug interaction statement.

### 3.10 Inter-model agreement

Across the 184 cells with an existing article, all three models returned the same code in 160 (87.0%): 98.2% for G6PD (56 of 57), 82.3% for teratogenicity (51 of 62), and 81.5% for QT (53 of 65) (Table 5). The 12 G6PD and teratogenicity disagreements were resolved by a pharmacist co-author; the 12 QT disagreements carry the majority code, examined in the sensitivity analysis above. Agreement was lowest for Arabic QT (mean pairwise κ 0.47).

**Table 5.**
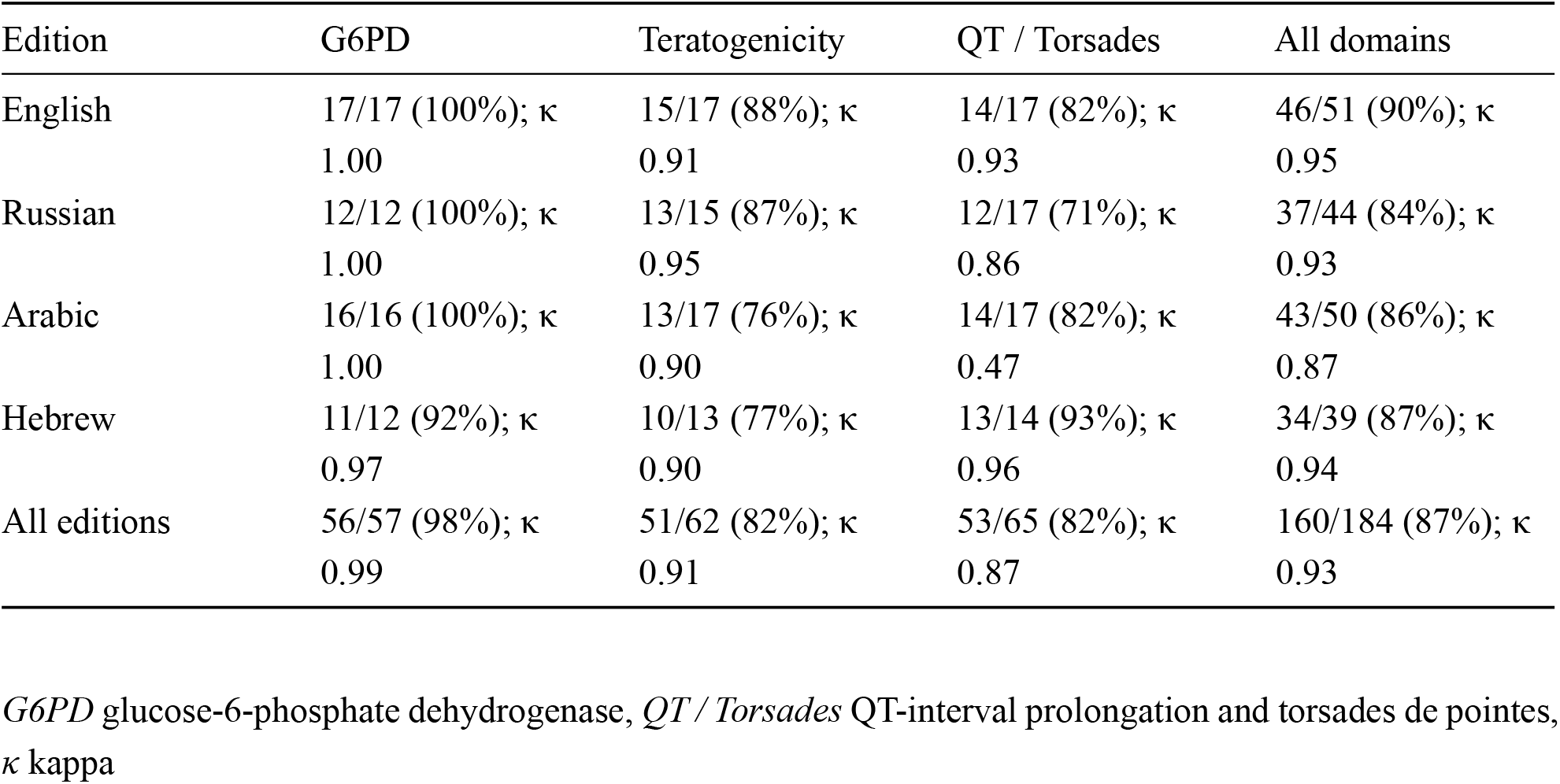
Inter-model agreement of the three classifiers by edition and domain, over cells with an existing article (risk-positive drugs and negative controls). Cells show models unanimous / cells (percent) and the mean pairwise quadratic-weighted κ. Agreement is computed from the model outputs and does not depend on any adjudication. Cells with no article are excluded because the models trivially return 0.

| Edition | G6PD | Teratogenicity | QT / Torsades | All domains |
| --- | --- | --- | --- | --- |
| English | 17/17 (100%); $\kappa$<br>1.00 | 15/17 (88%); $\kappa$<br>0.91 | 14/17 (82%); $\kappa$<br>0.93 | 46/51 (90%); $\kappa$<br>0.95 |
| Russian | 12/12 (100%); $\kappa$<br>1.00 | 13/15 (87%); $\kappa$<br>0.95 | 12/17 (71%); $\kappa$<br>0.86 | 37/44 (84%); $\kappa$<br>0.93 |
| Arabic | 16/16 (100%); $\kappa$<br>1.00 | 13/17 (76%); $\kappa$<br>0.90 | 14/17 (82%); $\kappa$<br>0.47 | 43/50 (86%); $\kappa$<br>0.87 |
| Hebrew | 11/12 (92%); $\kappa$<br>0.97 | 10/13 (77%); $\kappa$<br>0.90 | 13/14 (93%); $\kappa$<br>0.96 | 34/39 (87%); $\kappa$<br>0.94 |
| All editions | 56/57 (98%); $\kappa$<br>0.99 | 51/62 (82%); $\kappa$<br>0.91 | 53/65 (82%); $\kappa$<br>0.87 | 160/184 (87%); $\kappa$<br>0.93 |

*G6PD* glucose-6-phosphate dehydrogenase, *QT / Torsades* QT-interval prolongation and torsades de pointes, $\kappa$ kappa

### 3.11 Consistency checks of the instrument

These checks are indirect and are not a validation. Over the 85 risk-positive cells with an existing article, the three models were unanimous in 67 (78.8%): English 21 of 24 (87.5%), Hebrew 16 of 21 (76.2%), Arabic 18 of 23 (78.3%), and Russian 12 of 17 (70.6%) (`data/validation/precheck_inter_model_` `agreement.csv`). Unanimity was lower in every non-English edition than in English, so the instrument was less certain where the gaps were found (Limitations). On the 24 G6PD risk-positive cells with an article, the legacy keyword coder agreed exactly with the final code in 14 (58.3%) and never assigned a higher code; in the other 10 it assigned a lower one (`data/validation/precheck_llm_vs_keyword.csv`; Table S5 is a different comparison, over the original 40 G6PD cells). Because the keyword coder is known to miss mentions in morphologically rich languages, this check can only reveal mentions that the models missed, and it found none.

## 4 Discussion

The Russian, Arabic, and Hebrew editions of Wikipedia were less likely than English to give patients an actionable drug-safety warning in eight of nine language-by-domain comparisons, across three mechanistically unrelated safety domains; Russian QT, at parity with English, is the exception. These domains share no biological mechanism, so the recurrence suggests a property of the editions more than of any topic. The evidence is directional and imprecise: with 24 risk-positive drugs in the model the credible intervals are wide, and the joint probability that all nine estimates are below 1 is only 32% under the primary prior. The gap also falls hardest on the populations these editions serve. For example, G6PD deficiency is concentrated in exactly the Mediterranean, Middle Eastern, and North African communities the Arabic and Hebrew editions reach [35,36]. Although absolute readership in Hebrew and Arabic is smaller, the populations those editions principally serve carry substantially elevated G6PD deficiency prevalence [37,38,39], so an absent or unactionable warning in these editions is more consequential per reader, not less.

An alternative explanation is that non-English articles are simply shorter and less complete. This is an established finding [20]. It accounts for part of the gap, but not for the most interesting part.

Our exploratory decomposition suggests that the mechanism differs by edition. For Russian the gap looks like a coverage gap: the article is often missing or short, and once it mentions the risk it warns like English, although the interval for the existing-article estimate is wide. For Arabic the gap looks like an escalation gap and is not a length effect: even when the article discusses the risk, it is far less likely to escalate to an explicit instruction. Hebrew points the same way, though its interval reaching the boundary leaves the pattern suggestive rather than established. Even where a warning is present, the non-English editions tend to place it less prominently, in running text rather than in a dedicated safety section.

This mention-without-directive pattern is the study’s most specific exploratory finding and it is length-independent by construction. We report it as an observation and deliberately do not explain it. The audit codes what is on the page; editor motivation is not in the data, and any account of *why* one language edition behaves this way would be speculation. The design identifies the pattern; it cannot identify its cause. Article length lies on the causal path from edition to warning rather than confounding it. The total effect therefore remains the clinically relevant quantity: a patient reads the article that exists, whatever the reason it reads as it does.

Two implications follow. First, because Wikipedia is an upstream input to LLM pretraining, retrieval-augmented generation, and AI search summaries, a vernacular warning gap is a plausible candidate cause of the same gap in AI answers for those languages: an information-provenance problem, not only a Wikipedia problem. Second, Wikidata’s dedicated contraindication property is almost unused (12 statements across the knowledge base, none for the audited drugs), so populating it for risk-positive drugs is a concrete, low-cost intervention: a single structured edit can propagate to infobox-driven editions, a more scalable fix than editing prose edition by edition. We did not test whether such edits change article text or reader behaviour.

One could instead audit the AI systems directly, posing medication questions to chatbots in each language and scoring the answers. The two designs answer different questions, and auditing the source has three advantages. Article text is a fixed, public, versioned object, so the measurement is reproducible; chatbot output varies with prompt, session, and silently updated model versions. Auditing the source identifies where downstream gaps can originate, which an audit of outputs cannot show without provenance. And the source is editable: a documented gap in an article or in Wikidata points directly at a remediation, whereas a documented chatbot failure does not.

### 4.1 Limitations

The panel is small and purposive, covering four of roughly 300 language editions and 33 drugs, so the credible intervals are wide and the estimates do not extend to Wikipedia as a whole; the choice of editions is justified in the Introduction. The audit reads article text only, so infobox- and Wikidata-rendered content is not fully reconciled. The codes come from an imperfect instrument that was not validated against human coding: unanimous model codes were accepted as final without a systematic human recheck, a pharmacist resolved only the G6PD and teratogenicity disagreements, and the 12 QT disagreements carry the majority code, examined only by the extreme-code sensitivity analysis. The consistency checks (inter-model unanimity and the keyword baseline) do not measure accuracy, and unanimity was lower outside English (70.6% to 78.3% of risk-positive cells, against 87.5% in English), so instrument error may differ by language. Error that is the same in every language would bias the language contrasts toward 1, but systematic under-detection in the non-English editions could create or inflate a gap, and over-detection could hide one. A blinded validation, in which a pharmacist rater per language codes the risk-positive cells without seeing the model codes, is planned (Supplementary Methods). Wikipedia also changes continuously. Of the 54 existing articles stored in March, 25 had been edited by the 9/6/2026 snapshot (median change 0.2% of words, maximum 17%); the G6PD and teratogenicity codes of those articles were assigned on the earlier text, and drift was not measured for the 10 drugs coded on 29/4/2026, so content drift can add noise to the codes, and the archived revision identifiers fix the state analysed but not always the state coded. The most extreme cell magnitudes are prior-sensitive, though their direction is stable. The mention-to-directive and length-adjusted estimates rest on small conditional subsets, and edition–length collinearity limits what the adjusted model can identify. The exploratory layer, the finite-population summary, and the post hoc common-effect model were specified after the primary results were seen, and there is no preregistration. Several non-English control articles carried pregnancy-risk language English lacked; for those non-English deviations one untested possibility is that a language edition serves multiple regulatory jurisdictions, making language a coarse proxy for regulatory context: an avenue for future work, alongside extending the audit to lower-resource languages and piloting Wikidata remediation.

## 5 Conclusions

Patients who read about a medication on Wikipedia in Arabic, Hebrew, or Russian were less likely than English readers to meet an explicit safety instruction, in eight of nine language-by-domain comparisons across three unrelated classes of drug harm, although the estimates are imprecise. Exploratory analyses point to different mechanisms: in Russian the gap looks like a coverage gap (missing or short articles), and in Arabic an escalation gap, in which a risk is covered but not made actionable. As Wikipedia increasingly feeds AI systems that patients and clinicians consult, these cross-lingual gaps are candidate sources of downstream safety disparities. Wikidata’s contraindication property is almost unused, and populating it for risk-positive drugs is an immediate, low-cost place to intervene.

## Data Availability

Code, the master drug list, coded outputs, the LLM response log, the archived revision identifiers and drift, the model-reporting outputs (data/coded/hierarchical_reporting/), and the Wikidata audit are available in the project's public GitHub repository (https://github.com/bgbg/wikipedia-drug-warning-disparities). Raw fetched article text is excluded for size and licensing reasons but is reproducible from the published collection scripts and the recorded snapshot dates.

## Declarations

### Availability of data and material

Code, the master drug list, coded outputs, the LLM response log, the archived revision identifiers and drift (`data/snapshot_revisions.csv`, `data/snapshot_drift.csv`), the model-reporting outputs (`data/coded/hierarchical_reporting/`), and the Wikidata audit are available in the project’s public GitHub repository (https://github.com/bgbg/wikipedia-drug-warning-disparities). Raw fetched article text is excluded for size and licensing reasons but is reproducible from the published collection scripts and the recorded snapshot dates.

### Ethics approval

Not applicable. The study analysed only publicly available Wikipedia and Wikidata content and involved no human participants, their data, or biological material, so research ethics approval was not required.

### Consent to participate

Not applicable.

### Consent for publication

Not applicable.

### Code availability

The analysis code is in the same public GitHub repository (https://github.com/bgbg/wikipedia-drug-warning-disparities).

### Author contributions

Contributions follow the CRediT taxonomy.

- **Maya Berlin** (clinical pharmacist; co-lead). Conceptualization; Methodology (definition of the warning-coding scheme and its clinical thresholds); Investigation (drug-list selection and clinical approval; adjudication of the G6PD and teratogenicity cases where the language models disagreed); Formal analysis (clinical interpretation); Writing (review and editing).
- **Matitiahu Berkovitch** (teratologist and clinical toxicologist). Validation (clinical and scientific relevance of the drug list, the coding scheme, and the findings); Supervision (senior clinical oversight); Writing (review and editing).
- **Nofar Benassayag Kaduri** (clinical pharmacist). Investigation (clinical review of the drug list and the reference standards); Writing (review and editing).
- **Einat Gorelik** (clinical pharmacist and pharmacoepidemiologist). Investigation (clinical review of the drug list and the reference standards; review of the safety information and of the clinical questions framing the audit); Writing (review and editing).
- **Boris Gorelik** (data scientist; co-lead). Conceptualization; Methodology (study design; the three-model LLM-consensus classification protocol); Software (MediaWiki article collection and Wikidata audit pipeline; reproducibility); Formal analysis (ordinal Bayesian analysis and robustness checks); Visualization; Writing (original draft).

All authors reviewed and approved the final manuscript.

