## Supplementary Material for "Drug Safety Warnings in English, Russian, Arabic and Hebrew Wikipedia: A Cross-Sectional Audit of 33 Drugs Across Three Risk Domains"

15/10/2026

This file contains supplementary methods, tables, and figures referenced from the main text, followed by the technical appendices.

#### Supplementary Methods

**Statistical software and robustness battery.** The primary hierarchical cumulative-logit model was fitted in bambi 0.17 [1] on PyMC 5 [2] following standard ordinal-regression practice [3]. Robustness was assessed with (i) a frequentist cumulative-link model with drug-clustered standard errors [4], (ii) a prior-sensitivity sweep at prior SDs of 0.5, 1.5, and 5.0, (iii) a refit without the per-drug random effect, and (iv) a paired Wilcoxon signed-rank test; the clinician-facing effect size is Cliff's delta [5] with a drug-level bootstrap. Using LLMs as annotators with expert escalation follows evidence that they can match or exceed human annotators on text-classification tasks [6]. Full diagnostics (R-hat, effective sample size, divergences, posterior-predictive checks) are in Appendix B.

**Additional reporting.** There was no preregistration. The primary hierarchical model is prespecified; four additions are labelled in the main text and tabulated below (Tables S6 to S9). All are computed by `scripts/run_model_reporting.py` with the primary model's formula, priors, seed (23), and sampler settings, which the script first checks by refitting the primary model and comparing its pooled medians with the committed ones (exact match). (i) The finite-population summary is exp of the mean, over the three realised domains, of the population-level language effect plus the domain offset, beside the population-level exp of the language effect. (ii) The sensitivity table refits the primary model with a HalfNormal($\sigma$) prior on the across-domain SD ($\tau$) of the language effect, $\sigma = 1$ and $\sigma = 0.5$, beside the primary fit, whose default prior bambi sets to HalfNormal(5.77). (iii) The joint probabilities are the share of posterior draws in which every listed odds ratio is below 1. (iv) The post hoc common-effect model, `code ~ 1 + language + topic + (1 | drug)`, uses the same Normal(0, 1.5) prior on the language effects and the same sampler settings. In every estimate the 12 QT model disagreements carry the majority code, which is final for this submission.

**Planned validation (no results reported).** The codes have not been validated against human coding. A blinded validation is planned in which one pharmacist rater per language codes the 85 risk-positive cells with an existing article (English 24, Hebrew 21, Arabic 23, Russian 17) from the article text alone, without the model codes or the reference-standard label, using the same 0/1/2 rubric or "cannot tell". Its analysis rule was written on 3/10/2026 and is logged in `methodological_decisions.md`. The primary measure is the per-language quadratic-weighted kappa between rater and final code, pooled across domains, with a bootstrap 95% confidence interval (adequate at 0.70 or higher, strong at 0.80 or higher). Language-asymmetric under-detection is tested with an exact two-sided sign test on the rater minus final code differences ($p < 0.10$, final code below rater code), Arabic and Hebrew against English, and the impact on the main estimates is assessed by substituting rater codes. With a single rater per language, the validation will measure agreement with one rater, not with an independent reference standard.

#### Supplementary Tables

**Table S1.** Explicit (code-2) warnings among the eight risk-positive drugs per domain, by edition.

| Edition | G6PD (of 8) | Teratogenicity (of 8) | QT / Torsades (of 8) |
|---|---|---|---|
| English | 6 | 6 | 7 |
| Russian | 2 | 4 | 6 |
| Arabic | 2 | 4 | 1 |
| Hebrew | 4 | 4 | 2 |

**Table S2.** Warning-code composition (% of articles at each code), by edition and topic.

| Edition | Topic | % code 0 | % code 1 | % code 2 |
|---|---|---|---|---|
| English | G6PD | 12 | 12 | 75 |
| Russian | G6PD | 75 | 0 | 25 |
| Arabic | G6PD | 50 | 25 | 25 |
| Hebrew | G6PD | 25 | 25 | 50 |
| English | Teratogenicity | 0 | 25 | 75 |
| Russian | Teratogenicity | 25 | 25 | 50 |
| Arabic | Teratogenicity | 0 | 50 | 50 |
| Hebrew | Teratogenicity | 12 | 38 | 50 |
| English | QT / Torsades | 0 | 12 | 88 |
| Russian | QT / Torsades | 12 | 12 | 75 |
| Arabic | QT / Torsades | 62 | 25 | 12 |
| Hebrew | QT / Torsades | 50 | 25 | 25 |

**Table S3.** Unpooled per-cell cumulative odds ratios versus English (two original domains; secondary model; posterior mean and 95% credible interval, computed as the highest-density interval). The hierarchical model of Table 2 is the paper's primary estimate. Source: `data/coded/ordinal_analysis_four_topic_2dom/cumulative_ors.csv` and `prob_dominance.csv`.

| Edition | Topic | OR vs EN | 95% CrI | P(English warns more) |
|---|---|---|---|---|
| Russian | G6PD | 0.08 | [0.00, 0.25] | 99.8% |
| Arabic | G6PD | 0.26 | [0.01, 0.70] | 98.1% |
| Hebrew | G6PD | 0.51 | [0.01, 1.61] | 87.7% |
| Russian | Teratogenicity | 0.41 | [0.01, 1.32] | 91.5% |
| Arabic | Teratogenicity | 0.67 | [0.01, 2.09] | 81.7% |
| Hebrew | Teratogenicity | 0.49 | [0.01, 1.54] | 89.0% |

**Table S4.** Cliff's delta (English vs non-English; positive = English warns more strongly; CIs excluding 0 are firm disparities). Source: `data/coded/ordinal_analysis_four_topic_2dom/cliffs_delta.csv`.

| Comparison | Topic | Cliff's δ | 95% CI |
|---|---|---|---|
| EN vs RU | G6PD | +0.59 | [+0.25, +0.88] |
| EN vs AR | G6PD | +0.53 | [+0.16, +0.88] |
| EN vs HE | G6PD | +0.25 | [−0.19, +0.69] |
| EN vs RU | Teratogenicity | +0.31 | [+0.02, +0.69] |
| EN vs AR | Teratogenicity | +0.25 | [+0.00, +0.63] |
| EN vs HE | Teratogenicity | +0.28 | [+0.02, +0.63] |

**Table S5.** Keyword baseline versus LLM consensus on the original G6PD cells (agreement 30/40). Every disagreement is one-directional: the LLM consensus assigned the stronger code.

| Drug | Edition | Keyword code | LLM code |
|---|---|---|---|
| dapsone | English | 1 | 2 |
| dapsone | Russian | 0 | 2 |
| methylene blue | Arabic | 0 | 1 |
| methylene blue | Hebrew | 1 | 2 |
| nitrofurantoin | Arabic | 1 | 2 |
| nitrofurantoin | Russian | 0 | 2 |
| pegloticase | Hebrew | 1 | 2 |
| phenazopyridine | Hebrew | 1 | 2 |
| primaquine | Arabic | 1 | 2 |
| primaquine | Hebrew | 1 | 2 |

**Table S6.** Sensitivity of the pooled odds ratios to the prior on the across-domain SD ($\tau$) of the language effect. Pooled cumulative odds ratio versus English (posterior median [95% credible interval]), probability that English warns more, and posterior median of τ [95% credible interval]. Source: `data/coded/hierarchical_reporting/tau_prior_table.csv`.

| Prior on τ | Edition | Pooled OR [95% CrI] | P(English warns more) | τ [95% CrI] |
|---|---|---|---|---|
| HalfNormal(5.77), bambi default (primary) | Russian | 0.41 [0.01, 3.04] | 78% | 2.32 [0.01, 6.44] |
| HalfNormal(5.77), bambi default (primary) | Arabic | 0.25 [0.01, 2.10] | 87% | 1.99 [0.00, 6.16] |
| HalfNormal(5.77), bambi default (primary) | Hebrew | 0.31 [0.01, 2.15] | 87% | 1.41 [0.00, 5.17] |
| HalfNormal(1) | Russian | 0.34 [0.02, 1.44] | 90% | 1.13 [0.00, 2.21] |
| HalfNormal(1) | Arabic | 0.19 [0.01, 0.72] | 98% | 0.85 [0.00, 1.97] |
| HalfNormal(1) | Hebrew | 0.28 [0.03, 0.93] | 96% | 0.63 [0.00, 1.78] |
| HalfNormal(0.5) | Russian | 0.33 [0.04, 0.99] | 95% | 0.61 [0.00, 1.24] |
| HalfNormal(0.5) | Arabic | 0.17 [0.02, 0.48] | 100% | 0.42 [0.00, 1.09] |
| HalfNormal(0.5) | Hebrew | 0.28 [0.04, 0.76] | 98% | 0.36 [0.00, 0.97] |

**Table S7.** Joint posterior probabilities that the listed odds ratios versus English are all below 1, by prior on τ. Source: `data/coded/hierarchical_reporting/joint_posterior_probabilities.csv`.

| Event | Primary prior | HalfNormal(1) | HalfNormal(0.5) |
|---|---|---|---|
| All three pooled odds ratios below 1 | 59% | 84% | 94% |
| All nine language-by-domain odds ratios below 1 | 32% | 46% | 68% |
| The eight language-by-domain odds ratios other than Russian QT all below 1 | 67% | 79% | 88% |
| Russian: all three domain odds ratios below 1 | 45% | 55% | 74% |
| Arabic: all three domain odds ratios below 1 | 83% | 92% | 97% |

| Event | Primary prior | HalfNormal(1) | HalfNormal(0.5) |
|---|---|---|---|
| Hebrew: all three domain odds ratios below 1 | 81% | 88% | 94% |

**Table S8.** Finite-population summary (descriptive complement; the prespecified estimate is the population-level pooled odds ratio). Odds ratio versus English (posterior median [95% credible interval]). Source: `data/coded/hierarchical_reporting/finite_population_summary.csv`.

| Edition | Population-level pooled OR | Finite-population OR (mean of the three realised domain effects) | P(finite-population OR < 1) |
|---|---|---|---|
| Russian | 0.41 [0.01, 3.04] | 0.23 [0.02, 0.70] | 98.7% |
| Arabic | 0.25 [0.01, 2.10] | 0.12 [0.01, 0.32] | 100.0% |
| Hebrew | 0.31 [0.01, 2.15] | 0.21 [0.03, 0.58] | 99.5% |

**Table S9.** POST HOC common-effect model with domain fixed effects, `code ~ 1 + language + topic + (1 | drug)` (not prespecified). Odds ratio versus English (posterior median [95% credible interval]); the joint probability that all three odds ratios are below 1 is 98%. Convergence: R-hat 1.002, minimum bulk effective sample size 1,716, 0 divergences. Source: `data/coded/hierarchical_reporting/post_hoc_common_effect.csv`.

| Edition | OR [95% CrI] | P(English warns more) |
|---|---|---|
| Russian | 0.30 [0.06, 0.76] | 98.5% |
| Arabic | 0.16 [0.04, 0.40] | 100.0% |
| Hebrew | 0.25 [0.05, 0.61] | 99.4% |

### Supplementary Figures

### Appendix A. Technical and reproducibility details

#### A.1 Data access

Wikipedia articles were retrieved through the MediaWiki API. The analysed text is `data/raw/articles.jsonl`, which the repository history shows stored on 22/3/2026 (first 40 articles), 23/3/2026 (60 records), and 9/6/2026 (all 160 records, including the QT-domain drugs and a refresh of the earlier records); the 9/6/2026 file is the final snapshot used for the structural metrics. On 24/6/2026 its Hebrew reference counts were corrected (inline-note templates counted); the article text did not change. The articles of the 10 drugs added after the pharmacist review of the drug list were collected and classified for G6PD

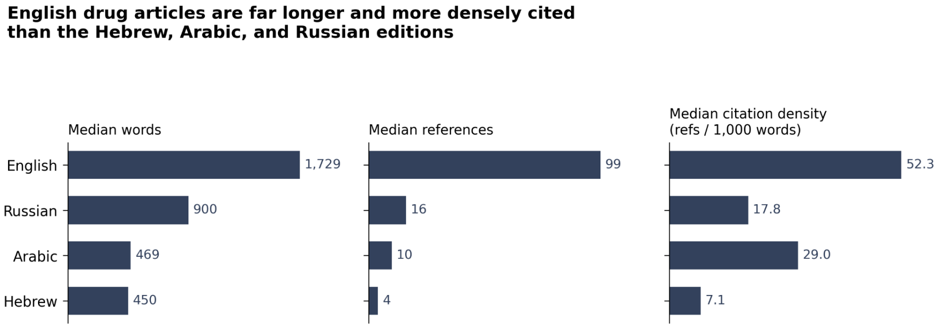


**Figure S1.** Structural quality by language (median word count, reference count, and citation density).

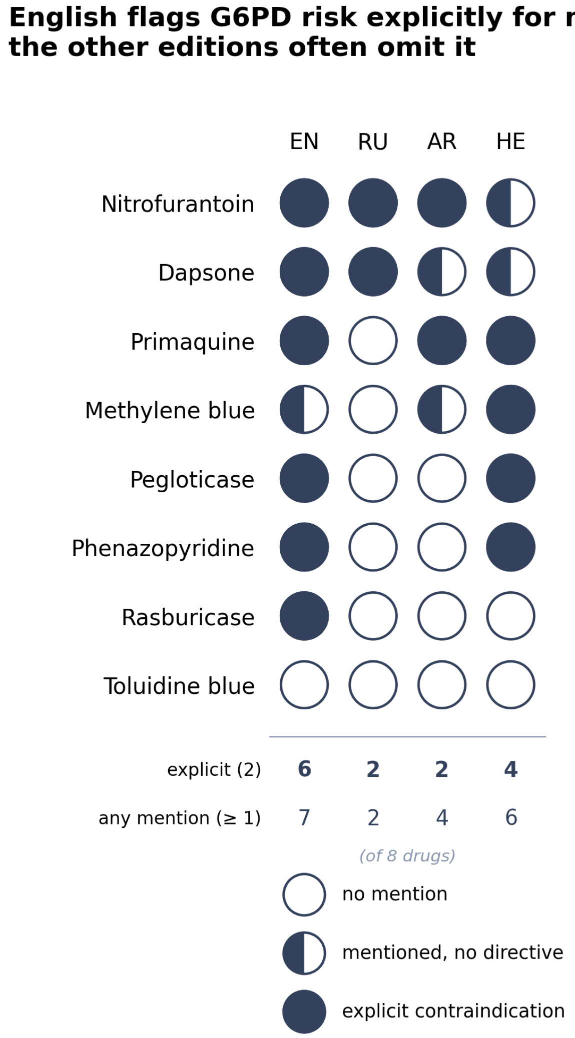


**Figure S2.** G6PD warning code by drug and edition.

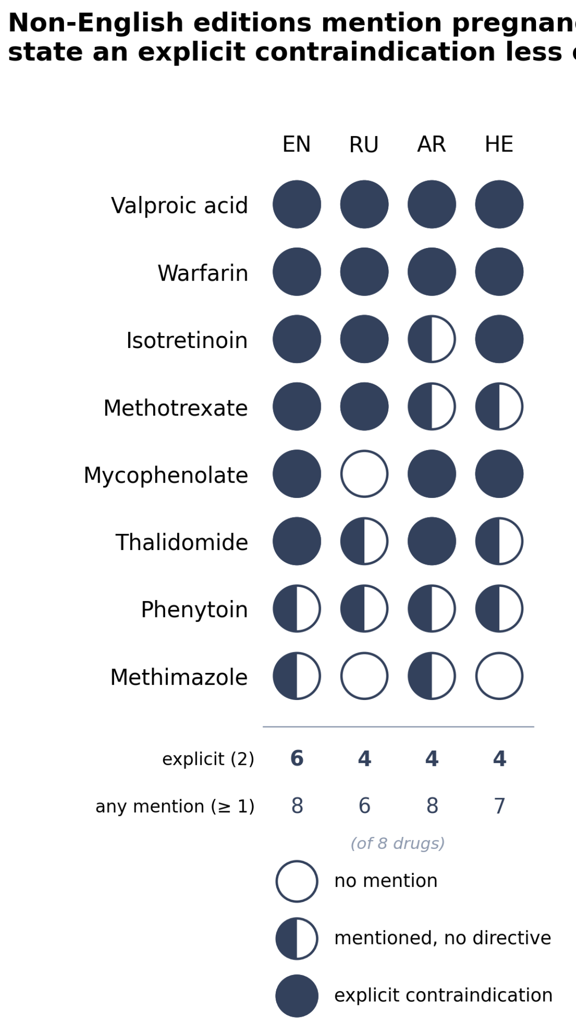


**Figure S3.** Teratogenicity warning code by drug and edition.

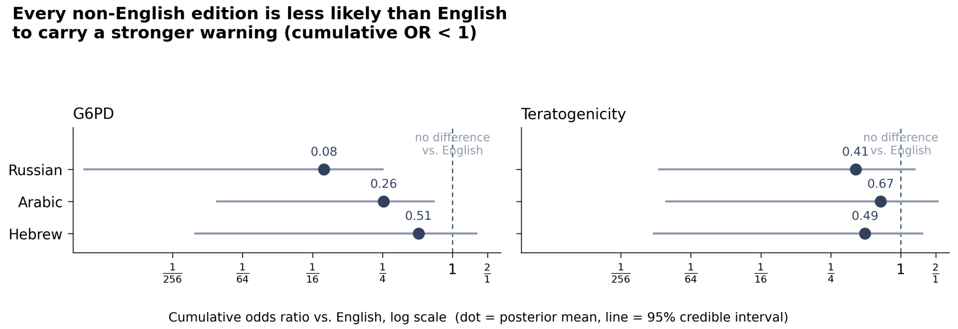


**Figure S4.** Per-cell cumulative odds ratios versus English, two original domains (secondary model; see Table S3).

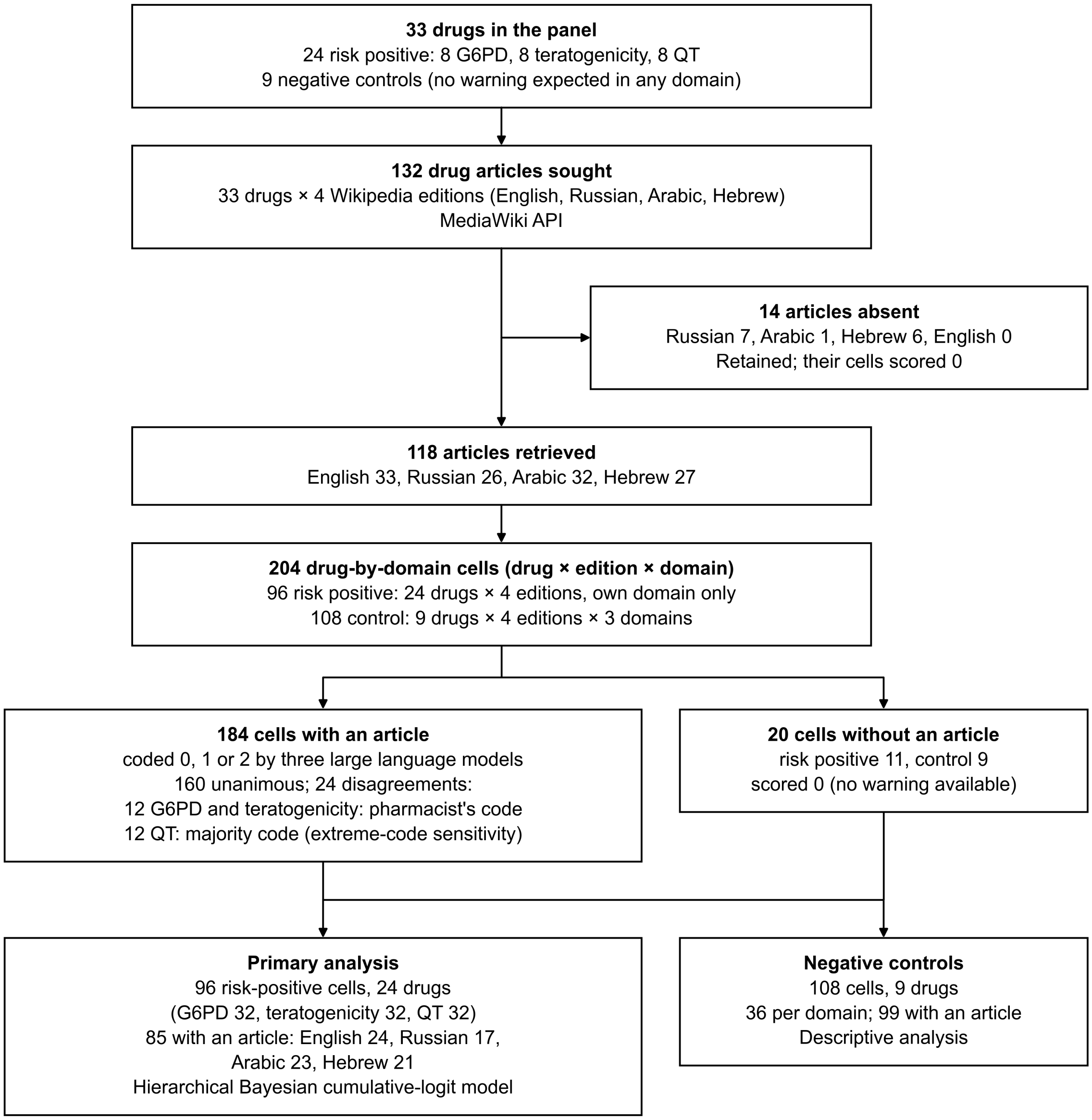


**Figure S5.** STROBE study flow diagram: drugs in the panel, Wikipedia articles sought and retrieved, drug-by-domain cells, coding, and the cells entering the primary analysis and the negative-control analysis. Absent articles were retained and their cells scored 0.

and teratogenicity on 29/4/2026 (commits `0178f22` and `d9d7f3d` in the project repository, on branch `develop/22-extend-dataset-to-25-drugs`, which also holds the model-call log of that run). The model calls therefore date from 23/3/2026 (G6PD and teratogenicity, first 15 drugs), 29/4/2026 (G6PD and teratogenicity, 10 later drugs), and 9/6/2026 (QT). An earlier version of this file dated the QT collection 14/6/2026; that date is not supported by the repository history and is replaced by 9/6/2026. The Wikidata structured-data audit was rerun on 2/10/2026 with the corrected properties. Pharmacist adjudication of the G6PD and teratogenicity disagreements was completed in May 2026; the 12 QT disagreements were not adjudicated and carry the majority code of the three models.

Because Wikipedia is mutable, the archived revisions define the article state analysed. For each of the 118 existing articles of the 33-drug panel, `scripts/fetch_revision_ids.py` queried the MediaWiki revision history for the revisions made on the stored last-edit date (UTC) and compared their wikitext with the stored text. All 118 matched a revision exactly (surrounding whitespace trimmed); the revision identifiers and timestamps are in `data/snapshot_revisions.csv` (last-edit dates 23/10/2022 to 9/6/2026) and can be opened with the `oldid` parameter. Content drift: of the 54 existing articles stored in March 2026, 25 differed in the final snapshot (median absolute change in word count 0.2%, maximum 17%, for isotretinoin in Russian), so the G6PD and teratogenicity codes of those articles were assigned on slightly earlier text than the archived revisions (`data/snapshot_drift.csv`, from `scripts/snapshot_drift.py`). Drift was not measured for the 10 drugs collected on 29/4/2026.

#### A.2 Classification models and decoding

The three consensus models were the OpenRouter endpoints openai/gpt-4o, google/gemini-2.5-flash, and x-ai/grok-3 for the G6PD and teratogenicity domains; for the later QT domain the third model was x-ai/grok-4.3 (Grok-3 having been deprecated; see Section 2.6, Coding instrument, of the main text). All were queried at temperature 0 with a domain-specific system prompt and few-shot worked examples (three for G6PD, six for teratogenicity, and three for QT); the full prompts are reproduced in Appendix D. Each call returned a JSON object (code, location, justification), and failed calls were retried once. Articles longer than 100,000 characters were truncated, and missing articles were assigned code 0 without a query.

#### A.3 Ordinal model and sampling

All ordinal models were fitted in bambi 0.17 [1] on PyMC 5 [2], following standard ordinal-regression practice [3]. The primary hierarchical model, code ~ 1 + language + (1 | drug) + (language | topic) with English as the reference edition, used a Normal(0, 1.5) prior on the language effects and bambi's default priors elsewhere; sampling used four chains of 1,500 draws after 1,500 tuning iterations at a target acceptance of 0.98 (seed 23), and its convergence diagnostics are reported in Results. The unpooled per-cell secondary model (Table S3), code ~ 0 + cell + (1 | drug) with cell the eight-level language-by-topic factor, used Normal(0, 1.5) priors on the cell effects and, for the drug random effect, a Normal effect whose standard deviation carried a HalfNormal(1) hyperprior; the cumulative thresholds used bambi's default ordered-Normal prior. Its sampling used four chains of 2,000 draws after 2,000 tuning iterations at a target acceptance of 0.95 (seed 23). The prior-sensitivity sweep refit the model at prior standard deviations of 0.5, 1.5, and 5.0, and the region of practical equivalence was set to odds ratios in [0.5, 2.0]. Robustness checks comprised a frequentist cumulative-link model (statsmodels) with drug-clustered standard errors [4], a refit without the random effect, and a paired Wilcoxon signed-rank test; Cliff's delta [5] used a drug-level bootstrap of 2,000 resamples. For the forest display in the main text (Fig. 3), 80% and 95% highest-density intervals were computed from the same posterior draws.

##### A.4 Software and availability

Analyses used Python 3 with bambi, PyMC, ArviZ, statsmodels, and SciPy. Code, the drug list, the coded outputs, the archived revision identifiers, the model-reporting outputs, and the Wikidata audit are available in the project repository; raw article text is excluded for size and licensing but is reproducible from the collection scripts and the archived revision identifiers above. The model-reporting script (`scripts/run_model_reporting.py`) documents a toolchain workaround for recent macOS versions; it does not touch the model.

#### Appendix B. Model diagnostics and robustness

The cross-lingual gaps reported in Results come from the Bayesian ordinal models of Appendix A.3; this appendix records the checks that justify relying on them.

**Convergence.** For the hierarchical primary model: $\hat{R}$ 1.003, minimum bulk effective sample size ≈ 1,435, and 0 divergences (reported in Results). For the refits of the reporting additions (`data/coded/hierarchical_reporting/convergence.csv`): HalfNormal(0.5) on $\tau$, $\hat{R}$ 1.002, minimum ESS 1,952, 0 divergences; HalfNormal(1), $\hat{R}$ 1.003, minimum ESS 1,606, 0 divergences; post hoc common-effect model, $\hat{R}$ 1.002, minimum ESS 1,716, 0 divergences. For the unpooled per-cell model: maximum R-hat 1.000 and minimum bulk effective sample size 2,351 across all parameters (`data/coded/ordinal_analysis_four_topic_2dom/manifest.json`).

**Frequentist robustness.** Refitting as a frequentist cumulative-link model (OrderedModel) with drug-clustered standard errors gives the same direction in all six cells:

| Edition | Topic | OR (Bayes) | OR (freq.) | 95% CI (freq.) |
|---|---|---|---|---|
| RU | G6PD | 0.08 | 0.04 | [0.00, 0.43] |
| AR | G6PD | 0.26 | 0.10 | [0.01, 0.90] |
| HE | G6PD | 0.51 | 0.33 | [0.04, 2.74] |
| RU | Teratogenicity | 0.41 | 0.28 | [0.07, 1.18] |
| AR | Teratogenicity | 0.67 | 0.42 | [0.11, 1.51] |
| HE | Teratogenicity | 0.49 | 0.35 | [0.11, 1.11] |

**Prior sensitivity.** Posterior-mean odds ratios vary up to 14.3× across the prior-SD grid {0.5, 1.5, 5.0} for the most extreme cells; the direction is stable across all priors, so magnitude statements (especially RU G6PD) are hedged.

**No random effect.** Refitting without (1 | drug) leaves every cell in the same direction and of similar magnitude (RU G6PD 0.08 → 0.11; RU teratogenicity 0.41 → 0.53; AR G6PD 0.26 → 0.34; AR teratogenicity 0.67

→ 0.74; HE G6PD 0.51 → 0.61; HE teratogenicity 0.49 → 0.64; `sensitivity_no_random_effect.csv`). The per-drug shrinkage does not produce the gap.

**Distributional cross-check** (paired Wilcoxon signed-rank, English vs. each non-English edition; 8 paired drugs):

| Edition | Topic | Non-zero pairs | p |
|---|---|---|---|
| RU | G6PD | 5 | 0.0625 |
| AR | G6PD | 4 | 0.125 |
| HE | G6PD | 4 | 0.500 |
| RU | Teratogenicity | 3 | 0.250 |
| AR | Teratogenicity | 2 | 0.500 |
| HE | Teratogenicity | 3 | 0.250 |

All point toward English carrying the stronger warning, but with only eight paired drugs none reaches conventional significance (smallest p = 0.0625, RU G6PD). This cross-check is consistent in direction with the model, not independent confirmation.

**Posterior-predictive check.** The model reproduces the observed code composition: 20 of the 24 observed proportions fall within the 95% posterior-predictive interval (under the analysis seed; a single boundary cell is sensitive to Monte-Carlo noise).

#### Appendix C. Drug list

The audit covers 33 drugs: eight risk-positive drugs in each of the three domains (24 in total) and nine shared negative controls. The original 25-drug panel (G6PD, teratogenicity, controls) was reviewed and approved by the pharmacist co-author (M.B.) on 29/4/2026; the QT domain was added on 9/6/2026 under the stated inclusion rules.

- **G6PD risk-positive drugs (8):** rasburicase, methylene blue, pegloticase, dapsone, nitrofurantoin, phenazopyridine, primaquine, toluidine blue. All are risk positive under the CPIC G6PD guideline, which grades them from “contraindicated” through “avoid in deficient patients” to “use with caution” [7].
- **Teratogenicity risk-positive drugs (8):** isotretinoin, valproic acid, methotrexate, warfarin, thalidomide, phenytoin, mycophenolate, methimazole. All are risk positive under FDA pregnancy labelling (REMS, black box, or PLLR narrative) [8,9].
- **QT / Torsades risk-positive drugs (8):** citalopram, azithromycin, levofloxacin, ondansetron, haloperidol, methadone, hydroxychloroquine, domperidone. Inclusion rule: drugs on the CredibleMeds *Known Risk of Torsades de Pointes* list (AZCERT, under an FDA Safe Use Initiative contract) that clear a lay-exposure bar, sampled across drug classes so the domain is not a single drug family. Domperidone, not approved in the United States but used across the EU, Israel, and the post-Soviet states, doubles as a probe of regulatory-jurisdiction variation.

- **Negative controls (9):** amoxicillin, atorvastatin, omeprazole, doxylamine, pyridoxine (B6), paracetamol, loratadine, famotidine, loperamide. Selected to have no known risk in any of the three domains; expected score 0 for every domain in all four languages. Documented exceptions are treated as data about Wikipedia, not coding errors: atorvastatin's FDA pregnancy contraindication was rescinded only in July 2021, so older revisions may still flag it as Category X; and the QT domain surfaces faithful code-1 mentions for loperamide and famotidine (drugs with QT signals at supratherapeutic doses), retained as controls under the *Known-Risk* (not *conditional-risk*) inclusion bar; the planned pharmacist sign-off of these controls was not obtained before submission.

#### Appendix D. Classification prompts

The verbatim system prompts and few-shot worked examples sent to the three models, reproduced from the versioned prompt file (`data/prompts/classification_prompts.json`). Each example is a user message (article text) followed by the expected assistant reply.

##### D.1 G6PD deficiency

System prompt:

```
You are a medical information classifier.
You will be given the full text of a Wikipedia drug article.
Your task is to determine whether the article contains a warning about G6PD
 ↪  (glucose-6-phosphate dehydrogenase) deficiency.

IMPORTANT RULES:
- The article may be written in ANY language, including English, Hebrew, Arabic,
 ↪  Russian, or others. You must classify the content accurately regardless of the
 ↪  language.
- Base your classification ONLY on the text provided. Do NOT use your own medical
 ↪  knowledge about the drug. If the article does not mention G6PD, classify as 0 even
 ↪  if you know the drug has a G6PD interaction.
- Only classify what is explicitly written in the article text.

Classify using this scale:
- 0: No mention of G6PD or G6PD deficiency anywhere in the article.
- 1: G6PD or G6PD deficiency is mentioned, but there is no explicit clinical action
 ↪  instruction (e.g., it appears in a pharmacology or mechanism section, or in a
 ↪  side-effects list without any directive to avoid the drug).
- 2: There is an explicit contraindication or avoidance instruction linked to G6PD
 ↪  deficiency. This includes ANY of the following:
    - The word 'contraindicated' used in connection with G6PD.
    - G6PD deficiency listed under a 'Contraindications' heading or section (in any
     ↪  language).
    - Phrases like 'avoid', 'do not use', 'should not be used' in connection with G6PD.
```

```
  The term 'contraindicated' is itself a clinical action instruction - if G6PD deficiency is listed as a contraindication, that is code 2 even without additional plain-language explanation.

Also identify WHERE in the article the warning appears:
- 'lead': in the introductory section before any headings
- 'contraindications_section': in a section about contraindications
- 'adverse_effects_section': in a section about adverse effects or side effects
- 'body': in another named section
- 'footnote_only': only in references/footnotes
- 'absent': no mention found

Respond with ONLY a JSON object in this exact format:
{"code": <0|1|2>, "location": "<location>", "justification": "<one sentence explaining your classification>"}
```

Few-shot examples (3):

Example 1.1, user message:

```
Classify this article for G6PD warnings:

Amoxicillin is a penicillin-type antibiotic used to treat bacterial infections. It works by stopping the growth of bacteria. Common side effects include nausea, vomiting, and diarrhea. It is generally considered safe during pregnancy.
```

Example 1.1, assistant reply:

```
{"code": 0, "location": "absent", "justification": "The article does not mention G6PD or glucose-6-phosphate dehydrogenase anywhere."}
```

Example 1.2, user message:

```
Classify this article for G6PD warnings:

Dapsone is a sulfone antibiotic used primarily for dermatitis herpetiformis and leprosy. It may cause hemolytic anemia, particularly in patients with glucose-6-phosphate dehydrogenase (G6PD) deficiency. Other side effects include methemoglobinemia and peripheral neuropathy.
```

Example 1.2, assistant reply:

```
{"code": 1, "location": "body", "justification": "G6PD deficiency is mentioned as a risk factor for hemolytic anemia, but no explicit avoidance instruction is given."}
```

Example 1.3, user message:

```
Classify this article for G6PD warnings:

Nitrofurantoin is an antibiotic used to treat urinary tract infections.

== Contraindications ==
```

```
Nitrofurantoin is contraindicated in patients with glucose-6-phosphate dehydrogenase
 ↪ (G6PD) deficiency due to the risk of hemolytic anemia. It should not be used in
 ↪ these patients.
```

Example 1.3, assistant reply:

```
{"code": 2, "location": "contraindications_section", "justification": "The
 ↪ contraindications section explicitly states the drug is contraindicated in
 ↪ G6PD-deficient patients."}
```

##### D.2 Teratogenicity

System prompt:

```
You are a medical information classifier.
You will be given the full text of a Wikipedia drug article.
Your task is to determine whether the article contains a warning about teratogenicity
 ↪ (risk of harm to a fetus during pregnancy).

IMPORTANT RULES:
- The article may be written in ANY language, including English, Hebrew, Arabic,
 ↪ Russian, or others. You must classify the content accurately regardless of the
 ↪ language.
- Base your classification ONLY on the text provided. Do NOT use your own medical
 ↪ knowledge about the drug. If the article does not mention pregnancy risk, classify
 ↪ as 0 even if you know the drug is teratogenic.
- Only classify what is explicitly written in the article text.

IMPORTANT - This study measures what information a NON-SPECIALIST reader (a patient, not
 ↪ a pharmacist) would get from the article. Code 2 requires a directive that a lay
 ↪ reader would understand as 'do not take this drug if pregnant.'

Classify using this scale:
- 0: No mention of pregnancy risk, teratogenicity, or fetal harm anywhere in the
 ↪ article. This ALSO includes:
    - FDA pregnancy risk categories A, B, or C appearing in an infobox WITHOUT any body
     ↪ text discussing pregnancy risk. These categories do not signal teratogenic risk.
    - Generic caution phrases like 'use with caution in pregnancy', 'consult your
     ↪ physician before using during pregnancy', or 'should be used only if clearly
     ↪ needed' - these are NOT teratogenicity warnings.
- 1: Pregnancy risk or teratogenicity is mentioned, but there is no explicit avoidance
 ↪ instruction. This includes:
    - Teratogenic effects described factually (e.g., 'causes birth defects', 'associated
     ↪ with fetal malformations') WITHOUT a directive to avoid the drug.
    - Historical or narrative descriptions of a drug's teratogenic effects (e.g.,
     ↪ describing a thalidomide-like disaster, or explaining the mechanism of fetal
     ↪ harm) WITHOUT a clinical directive to avoid the drug in pregnancy. Even if the
     ↪ entire article is about the drug being a teratogen, if there is no explicit
     ↪ 'avoid in pregnancy' or 'contraindicated in pregnancy' statement, this is code 1.
```

```
    - FDA pregnancy risk categories D or X appearing ONLY in an infobox, without body
     ↪  text explaining the risk or giving an avoidance directive. The letter alone is a
     ↪  technical code that a lay reader cannot act on. Do NOT code infobox-only D or X
     ↪  as code 2.
    - Pregnancy risk discussed in a pharmacology or mechanism section without clinical
     ↪  guidance.
- 2: There is an explicit contraindication in pregnancy or instruction to avoid the drug
 ↪  during pregnancy that a lay reader would understand. This includes ANY of the
 ↪  following:
    - The word 'contraindicated' used in connection with pregnancy.
    - Pregnancy listed under a 'Contraindications' heading or section (in any language).
    - Phrases like 'avoid in pregnancy', 'must not be used by pregnant women', 'do not
     ↪  take if pregnant', 'women of childbearing potential must use contraception'.
  The term 'contraindicated' is itself a clinical action instruction - if pregnancy is
   ↪  listed as a contraindication, that is code 2 even without additional
   ↪  plain-language explanation.
  NOTE: If body text contains an explicit directive, the body text determines the code
   ↪  regardless of what the infobox says.

Also identify WHERE in the article the warning appears:
- 'lead': in the introductory section before any headings
- 'contraindications_section': in a section about contraindications
- 'adverse_effects_section': in a section about adverse effects or side effects
- 'body': in another named section
- 'footnote_only': only in references/footnotes
- 'absent': no mention found

Respond with ONLY a JSON object in this exact format:
{"code": <0|1|2>, "location": "<location>", "justification": "<one sentence explaining
 ↪  your classification>"}
```

Few-shot examples (6):

Example 2.1, user message:

```
Classify this article for teratogenicity warnings:

Amoxicillin is a penicillin-type antibiotic used to treat bacterial infections. It works
 ↪  by stopping the growth of bacteria. Common side effects include nausea, vomiting,
 ↪  and diarrhea. It is generally considered safe during pregnancy.
```

Example 2.1, assistant reply:

```
{"code": 0, "location": "absent", "justification": "The article mentions pregnancy but
 ↪  states the drug is safe, with no teratogenicity warning."}
```

Example 2.2, user message:

```
Classify this article for teratogenicity warnings:
```

```
Warfarin is an anticoagulant medication. It is used to prevent blood clots. Warfarin
↪ crosses the placenta and has been associated with birth defects when used during
↪ pregnancy, particularly during the first trimester. Fetal effects include nasal
↪ hypoplasia and skeletal abnormalities.
```

Example 2.2, assistant reply:

```
{"code": 1, "location": "body", "justification": "Teratogenic effects are described
↪ (birth defects, nasal hypoplasia) but no explicit contraindication or avoidance
↪ instruction is stated."}
```

Example 2.3, user message:

```
Classify this article for teratogenicity warnings:

{{Infobox drug
| pregnancy_AU = B
| pregnancy_US = B
}}
Amoxicillin is a broad-spectrum antibiotic. It is commonly used to treat respiratory and
↪ urinary tract infections. The drug penetrates the placenta and is excreted in breast
↪ milk. Caution is advised for pregnant or lactating women; use only under medical
↪ supervision.
```

Example 2.3, assistant reply:

```
{"code": 0, "location": "absent", "justification": "Infobox category B does not signal
↪ teratogenic risk. The body text only contains generic caution language ('caution is
↪ advised', 'use under medical supervision') which is not a teratogenicity warning."}
```

Example 2.4, user message:

```
Classify this article for teratogenicity warnings:

{{Infobox drug
| pregnancy_AU = D
| pregnancy_US = X
}}
Methotrexate is a chemotherapy agent and immune system suppressant. It is used to treat
↪ certain types of cancer, rheumatoid arthritis, and psoriasis. Common side effects
↪ include nausea, fatigue, and increased risk of infection.
```

Example 2.4, assistant reply:

```
{"code": 1, "location": "lead", "justification": "The infobox contains pregnancy risk
↪ categories D (AU) and X (US) which signal real risk, but the article body does not
↪ discuss pregnancy at all. The infobox letter alone is code 1, not code 2."}
```

Example 2.5, user message:

```
Classify this article for teratogenicity warnings:
```

```
Thalidomide is a sedative drug that became widely known for its teratogenicity. In the
↪ late 1950s and early 1960s, thousands of children were born with severe birth
↪ defects including phocomelia (shortened or absent limbs) after their mothers took
↪ thalidomide during pregnancy. The drug was withdrawn from most markets in 1961. The
↪ thalidomide disaster led to major reforms in drug testing regulations worldwide.
```

Example 2.5, assistant reply:

```
{"code": 1, "location": "body", "justification": "The article extensively describes the
↪ historical teratogenic disaster and birth defects caused by thalidomide, but
↪ contains no explicit directive to avoid the drug in pregnancy - it reads as a
↪ historical narrative, not a clinical warning."}
```

Example 2.6, user message:

```
Classify this article for teratogenicity warnings:

Isotretinoin is a retinoid used for severe acne.

== Contraindications ==
Isotretinoin is absolutely contraindicated in pregnancy. It is a known teratogen that
↪ causes severe birth defects including craniofacial, cardiac, and central nervous
↪ system malformations. Women of childbearing potential must use two forms of
↪ contraception and have negative pregnancy tests before starting treatment.
```

Example 2.6, assistant reply:

```
{"code": 2, "location": "contraindications_section", "justification": "The
↪ contraindications section explicitly states the drug is contraindicated in pregnancy
↪ and describes it as a known teratogen with mandatory pregnancy prevention measures."}
```

#### D.3 QT prolongation / Torsades de Pointes

System prompt:

```
You are a medical information classifier.
You will be given the full text of a Wikipedia drug article.
Your task is to determine whether the article contains a warning that the drug can
↪ prolong the QT interval or cause Torsades de Pointes (a dangerous ventricular
↪ heart-rhythm disturbance).

IMPORTANT RULES:
- The article may be written in ANY language, including English, Hebrew, Arabic,
↪ Russian, or others. You must classify the content accurately regardless of the
↪ language.
- Base your classification ONLY on the text provided. Do NOT use your own medical
↪ knowledge about the drug. If the article does not mention the risk, classify as 0
↪ even if you know the drug carries it.
- Only classify what is explicitly written in the article text.

Classify using this scale:
```

```
- 0: No mention of QT prolongation, the QT interval, Torsades de Pointes, or a drug-induced dangerous ventricular arrhythmia / heart-rhythm disturbance anywhere in the article.
- 1: QT prolongation, Torsades, or arrhythmia risk is mentioned, but there is no explicit clinical action instruction (e.g., it appears in a pharmacology/mechanism section, or in a side-effects list without any directive).
- 2: There is an explicit directive linked to QT/Torsades risk. This includes ANY of the following:
    - The word 'contraindicated' used in connection with QT prolongation, long-QT syndrome, or co-administration with other QT-prolonging drugs.
    - QT prolongation or 'risk of Torsades' listed under a 'Contraindications' or 'Warnings' heading with an avoidance directive (in any language).
    - Instructions such as 'avoid', 'do not use', 'should not be used', 'use only with ECG monitoring', or 'avoid in congenital long-QT syndrome / with low potassium or magnesium' in connection with the cardiac risk.
  The term 'contraindicated' is itself a clinical action instruction.

Also identify WHERE in the article the warning appears:
- 'lead': in the introductory section before any headings
- 'contraindications_section': in a section about contraindications
- 'adverse_effects_section': in a section about adverse effects or side effects
- 'body': in another named section
- 'footnote_only': only in references/footnotes
- 'absent': no mention found

Respond with ONLY a JSON object in this exact format:
{"code": <0|1|2>, "location": "<location>", "justification": "<one sentence explaining your classification>"}
```

Few-shot examples (3):

Example 3.1, user message:

```
Classify this article for QT-prolongation / Torsades de Pointes warnings:

Amoxicillin is a penicillin-type antibiotic used to treat bacterial infections. Common side effects include nausea, diarrhea, and rash.
```

Example 3.1, assistant reply:

```
{"code": 0, "location": "absent", "justification": "The article does not mention QT prolongation, Torsades, or any cardiac rhythm risk."}
```

Example 3.2, user message:

```
Classify this article for QT-prolongation / Torsades de Pointes warnings:

Azithromycin is a macrolide antibiotic. == Side effects == Reported adverse effects include diarrhea, nausea, and prolongation of the QT interval.
```

Example 3.2, assistant reply:

```
{"code": 1, "location": "adverse_effects_section", "justification": "QT prolongation is
↪ listed as a side effect but the article gives no directive to avoid or monitor."}
```

Example 3.3, user message:

```
Classify this article for QT-prolongation / Torsades de Pointes warnings:

Citalopram is an antidepressant. == Contraindications == Citalopram is contraindicated
↪ in patients with congenital long QT syndrome and should not be combined with other
↪ QT-prolonging medications because of the risk of Torsades de Pointes.
```

Example 3.3, assistant reply:

```
{"code": 2, "location": "contraindications_section", "justification": "The
↪ contraindications section explicitly bars use in long-QT patients and with other
↪ QT-prolonging drugs due to Torsades risk."}
```
